# COVID-19 Vaccine: Newspaper Coverage of the side effects of the vaccine in Nigeria

**DOI:** 10.1101/2021.10.02.21264454

**Authors:** Soyemi Kehinde Victor, Olagoke Ewedairo, Charles Olomofe

## Abstract

**Background:** COVID-19 Vaccine hesitancy is increasing globally, and this threatens the world’s ability to bring the pandemic under control. The way the media reports on the vaccine may influence or affect how the population perceive the safety and efficacy of the vaccine.

**Methods:** The aim of this study was to determine how newspapers in Nigeria report stories about the vaccine and the side effects of the vaccine amidst the growing fear on the safety of the vaccine. A total of 4 national daily newspapers were randomly selected for the study. These are *Leadership, Guardian, Nation and Punch* newspapers. The study was anchored on agenda setting theory. Quantitative content analysis research was used for the study. The duration of the study was the day the vaccine was introduced in Nigeria: March 1^st^, 2021 to July 31^st^, 2021. An Excel sheet served as the instrument for data collection and analysis done using SPSS version 25 with the level of significance predetermined at a p-value <0.05.

**Results:** Key findings from this research were: Government officials and technical experts were predominantly used by the newspapers as the source of their information. There was a mixed reporting of vaccine side effects with a significant difference between those newspaper publications that reported vaccine side effects and those that didn’t. Amongst those that reported side effects, there was also a significant difference between those that communicated how and where to report the side effects as against those that didn’t.

**Conclusion:** As part of the effort to curtail vaccine hesitancy, a continuous improvement in communicating the vaccine efficacy and safety is needed.

## BACKGROUND

### COVID-19, Nigeria and Covid-19 Vaccines

Corona viruses are part of a large family of viruses that are known to cause illnesses ranging from mild common cold to a more severe diseases such as Middle East Respiratory Syndrome (MERS) and Severe Acute Respiratory Syndrome (SARS).^1^In December 2019, in Wuhan Community in China, a rapidly infectious respiratory disease caused by a novel virus belonging to the Coronaviridae family was identified and appropriately named Severe Acute Respiratory Syndrome Corona virus-2 (SARS-CoV-2) and the disease called coronavirus disease 2019 (COVID-19). Due to a global increase in spread of COVID-19 and it’s attending risk, the World Health Organization (WHO) declared it a Public Health Emergency of International Concern (PHEIC) towards the end of January 2020.^2^ The first index case of the disease in Nigeria was reported in Lagos state in Feb 27^th^, 2020 by a foreigner who had come in from one of the high burden state^3^. As at 4^th^ July 2021: Total confirmed cases of COVID-19 in Nigeria stood at 167859 cases with 164382 discharged and 2121 deaths across the various states including the Federal Capital Territory^4^.

Several measures have been advocated for curtailing the spread of this pandemic; From good respiratory hygiene, social distancing, use of face mask and face shield, good respiratory hygiene and hand washing or sanitizing to now the need for Vaccination.^5^

Vaccine development takes significant years for it to pass through all the various stages before its approval for clinical use but in the prevailing circumstances that COVID-19 pose as a global pandemic on every aspect of life of human series of vaccines needed to be given emergency use listing by WHO (EUL) : Pfizer/BioNtech Comirnaty vaccine (EUL) on 31 December 2020, AstraZeneca/Oxford vaccine (EUL) on 16 February 2021, the Janssen vaccine (EUL) on 12 March 2021, Moderna COVID-19 vaccine was (EUL) on 30 April 2021, Sinopharm COVID-19 vaccine (EUL) on 7 May 2021 and the Sinovac-CoronaVac was listed for EUL on 1 June 2021.^6^

Nigeria received its first batch 3.94million doses of AstraZeneca/Oxford vaccine in March 2021 amidst a large media coverage with the president; Muhammed Buhari receiving his first dose on March 6^th^, 2021^7,8^. As at July 3^rd^, 2021, only about 1.34 Million people had received the 2 doses of the vaccine representing 0.6% of the country population with Lagos state accounting for a larger percentage of the vaccinated population and Bayelsa state with the least number of fully vaccinated population^9^.

### Media: COVID-19 related news and vaccine coverage

The role of the media in reporting health related information especially during pandemic is very important not only in keeping the general population informed but also helping to allay fears that usually hover around during pandemics^10,11^. Extensive media coverage during pandemics help curtail the panic and spread of rumor that are often associated with disease outbreaks especially a pandemic such as COVID-19. In a study carried out by Olapegba et.al in Nigeria to assess the knowledge of the populace on COVID-19^12^, the mass media (Television, radio and Newspaper) provided the most source of ‘go-to’ for information. In another study carried out to assess the knowledge and perceptions and attitude of Egyptians towards COVID-19^13^, knowledge, majority of the respondents had good general knowledge about the mode of transmission and prevention of the disease with a strong influence of their awareness due to a massive media-driven campaign on Mass media and Social media. Level of knowledge among rural dwellers was lower when compared with Urban dwellers showing the level of penetration of the media.

Vaccine hesitancy and uptake remains a global challenge, but worse hit is in developing countries. Vaccine refusal can lead to disease outbreaks with a typical example seen during the Polio outbreaks in Northern Nigeria in 2003 leading to a significant rise in Polio incidence in the country^14^. This refusal was sparked by rumors that vaccines were unsafe and aimed at controlling the sterility of a religious sect as well as spreading HIV.

**Fig 1:**
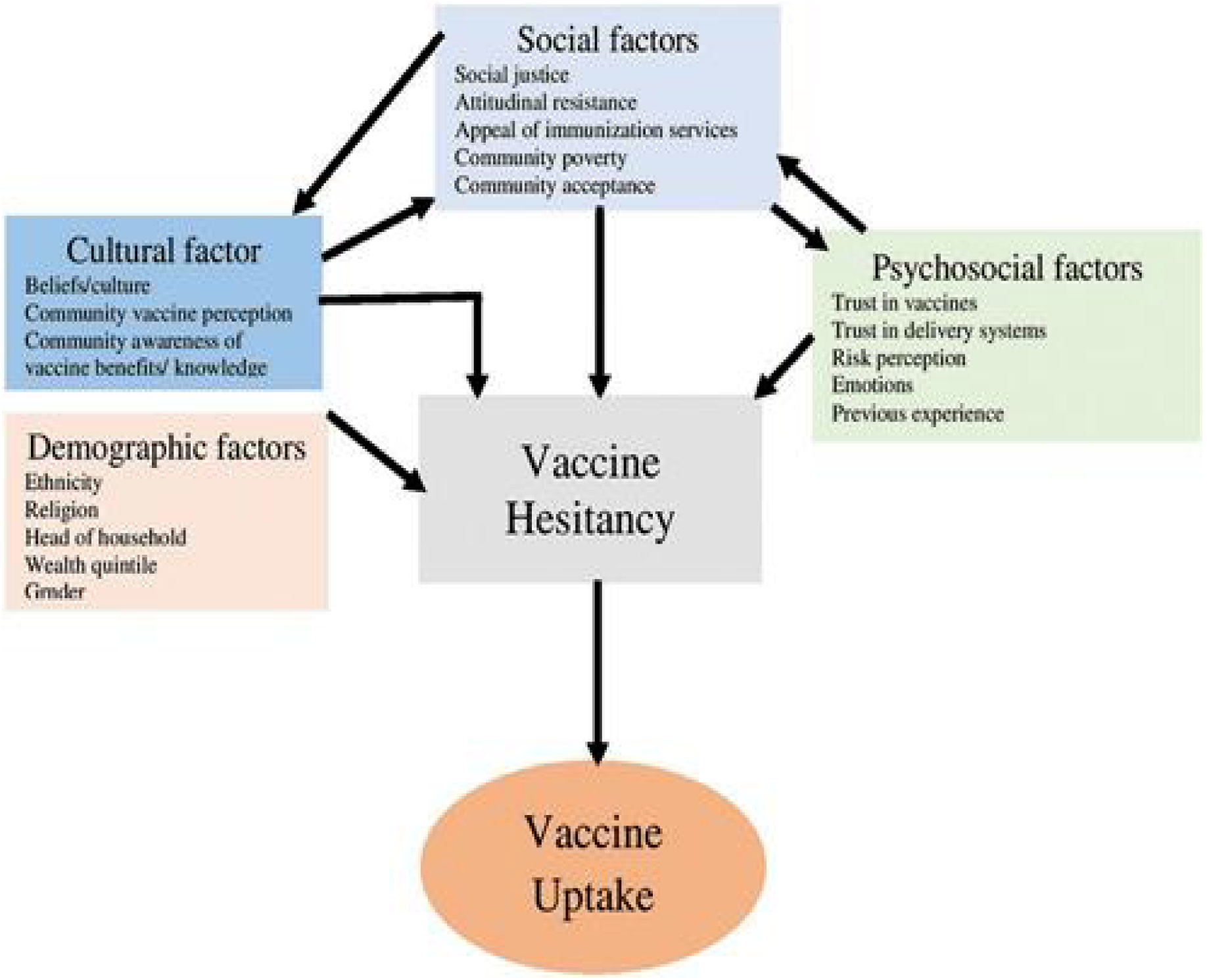
A conceptual framework on vaccine hesitancy. Figure from: Ogundele, Olorunfemi & Ogundele, Tolulope & Beloved, Omolola. (2020). Vaccine hesitancy in Nigeria: Contributing factors – way forward. The Nigerian Journal of General Practice. 18. 1. 10.4103/NJGP.NJGP_28_19.

The fear of the unknown side effects of the vaccine and the media projections of the side effect of the vaccine might also contribute to the vaccine hesitancy. An International media channel reported this’ A greater number of [heart inflammation cases](https://www.foxnews.com/health/myocarditis-signs-symptoms)_cropped up among members of the U.S. military than expected following mRNA [COVID-19 vaccination](https://www.foxnews.com/category/health/infectious-disease/coronavirus), a study found, though authors emphasized the benefits of the shots exceed risks of rare adverse events^15,16.^List of side effects of the Astrazeneca/Oxford Vaccine (Vaxzervia) listed in the Prescribing information leaflets showed pain at Injection site, injection site tenderness, headaches, myalgia, malaise as well as fever to be the most common side effects while thrombocytopenia is also said to be common.^17^. The bleeding tendency following the vaccine administration has pose a significant challenge as the information about the safety of the vaccines begins to unfold.

Reporting adverse events (Side effects) of a medication including vaccines help in the post-marketing surveillance for the pharmaceutical companies as well as the Drug regulatory bodies. In Nigeria the National Agency for Food and Drug Control established in October 1992 is saddled with regulation and control for those manufacturing, importing, exporting, distributing, and those selling and consuming Drugs, medical devices as well as vaccines^18^. NAFDAC has a Med Safety App^19^ where adverse events for drugs and including the COVID-19 vaccines can be reported by the general populace. The question is how much of this information is available to the public and how much has the mass media helped in projecting this information aside the media mention of the side effects of the vaccines which might contribute to the vaccine hesitancy in Nigeria

This study is needed to look at how newspaper report the side effects of COVID-19 Vaccines as a way of improving the vaccine uptake and allaying the fears among Nigerians.

### Objectives of the Study

The main objective of this study was to determine how the media report COVID-19 Vaccine and related side effects in Nigeria. The specific objectives of the study are:

1. To determine the sources cited in newspaper coverage on COVID-19 Vaccine.
2. To determine if newspaper talk on COVID-19 vaccine side effects.
3. To determine the story type which the newspapers use to report stories concerning the Vaccine side effects.
4. To ascertain if newspaper stories make suggestions regarding how to report the side effects of the vaccines

## METHODS

This is a descriptive quantitative content analysis carried out by sampling recognised newspapers with online presence and wide coverage in Nigeria with study period starting from March 2^nd^, 2021 when Nigeria received the first batch of the vaccine to July 2021 when the first phase of immunization cycle was stopped.

### Technique and Criteria for selecting stories

Use of keywords ‘COVID-19’ ‘Coronavirus’ ‘COVID-19 vaccines’ ‘Astrazeneca Vaccine’ ‘Vaccine side effects’ ‘COVID-19 vaccine side effect’ ‘Immunization against COVID-19’ ‘Death from COVID-19 vaccines’ ‘Bleeding from COVID-19’ ‘Coronavirus vaccine’

### Inclusion Criteria

- News mentioning COVID-19 Vaccine in Nigeria
- News on COVID-19 Vaccine side effects in Nigeria

### Exclusion Criteria

- News mentioning COVID-19 Vaccines with side effects but from other countries
- News mentioning only COVID-19 without the vaccines were excluded
- Articles with inadequate data related to the topic were also excluded

**Fig 2:**
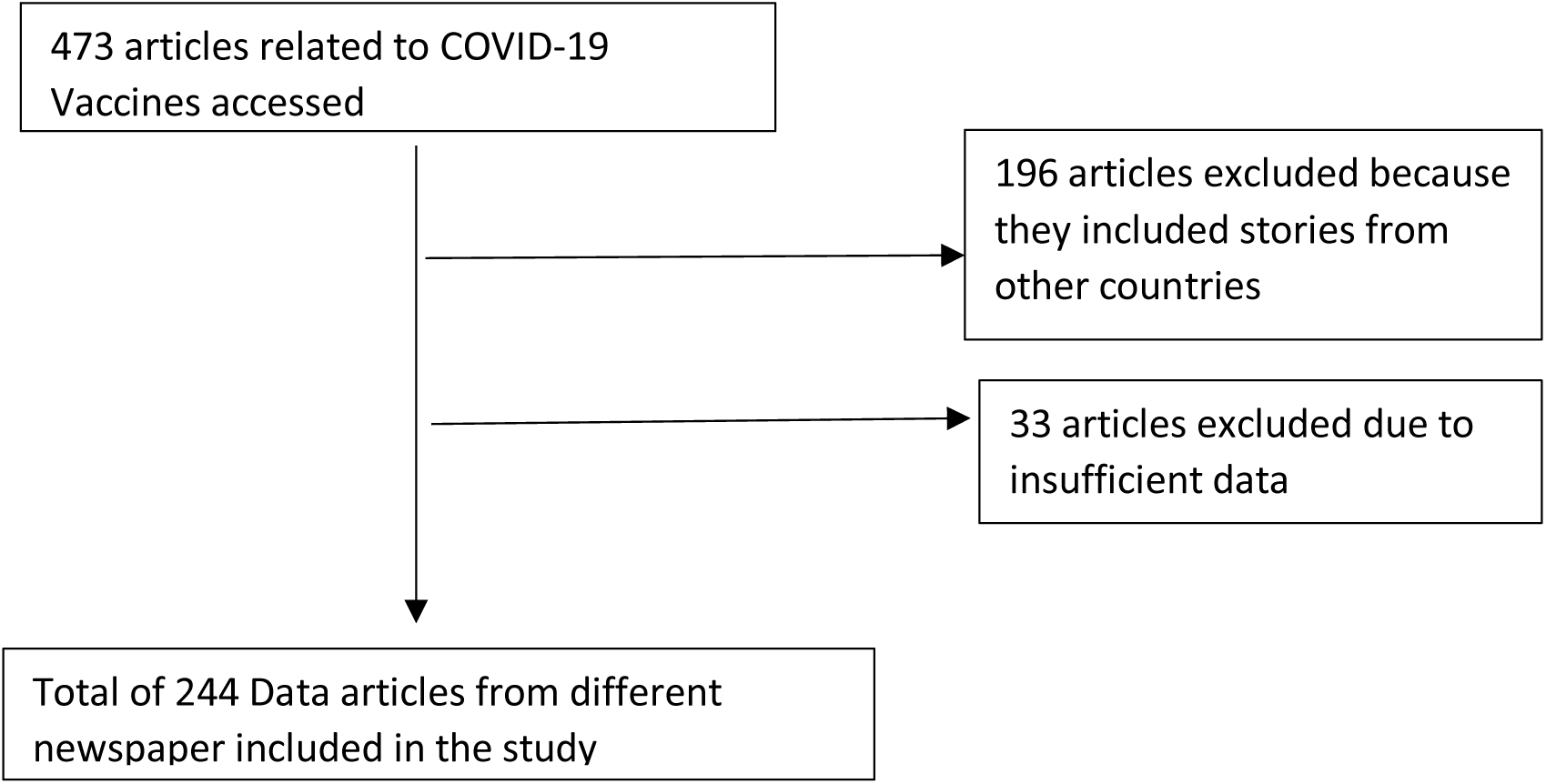
Article Selection flow chart.

### Unit of Analysis and Coding

The following highlighted how the information were coded and analysed

#### Newspaper

Newspaper in this case referred to the 4-newspaper analysed: The Guardian, The Nation, Punch and The Leadership. These newspapers were selected because of their National reach as well as their online presence.

#### Article type

The articles were classified based on the newspaper categorisation of the article into: News, Editorials, Opinion, Columns, Features and Others (Those without a classification heading)

#### Sources

The sources of information were classified under the following headings:

i. Government Officials and political office holders’ examples: Governors, Presidency
ii. Technical experts including healthcare practitioners and health related agencies
iii. Non-governmental and Religious Organizations not related to health industries
iv. Victims/Recipients of the Vaccine telling their stories or interviewed by the newspaper
v. Others including journalist write-up on the topic or where no source to the information could not be traced.

#### Side effects of vaccines mentioned

*The side effects the vaccine mentioned were coded as follows:*

i. None mentioned
ii. Fever
iii. Pain
iv. Bleeding
v. Myalgia/body pains
vi. Chills/cold
vii. Mixed (More than 1 side effects mentioned/unspecific)

#### Where to report the vaccine side-effects

i. Those that mentioned where to report represented as YES
ii. Those that did not mention where to report represented as NO

#### Data Capture and Analysis

Data captured on Microsoft excel sheet and analysis done with SPSS version 25. Descriptive statistics involving frequencies and percentages were run to describe article characteristics as well as to give a good understandable view of the information. Chi-square analysis with p-value set at 0.05 significance were done to analyse the relevance of the data.

## RESULTS

Below shows the results of the frequency distribution and analysis of the data 244 samples(n=244) were analysed across the newspaper after appropriately excluding the ones that were not relevant to the study (N=473). The Guardian Newspaper was responsible for 37.7% (n=92) while Punch newspaper had the least publication of 16.4% (n=40).

**Figure 3:**
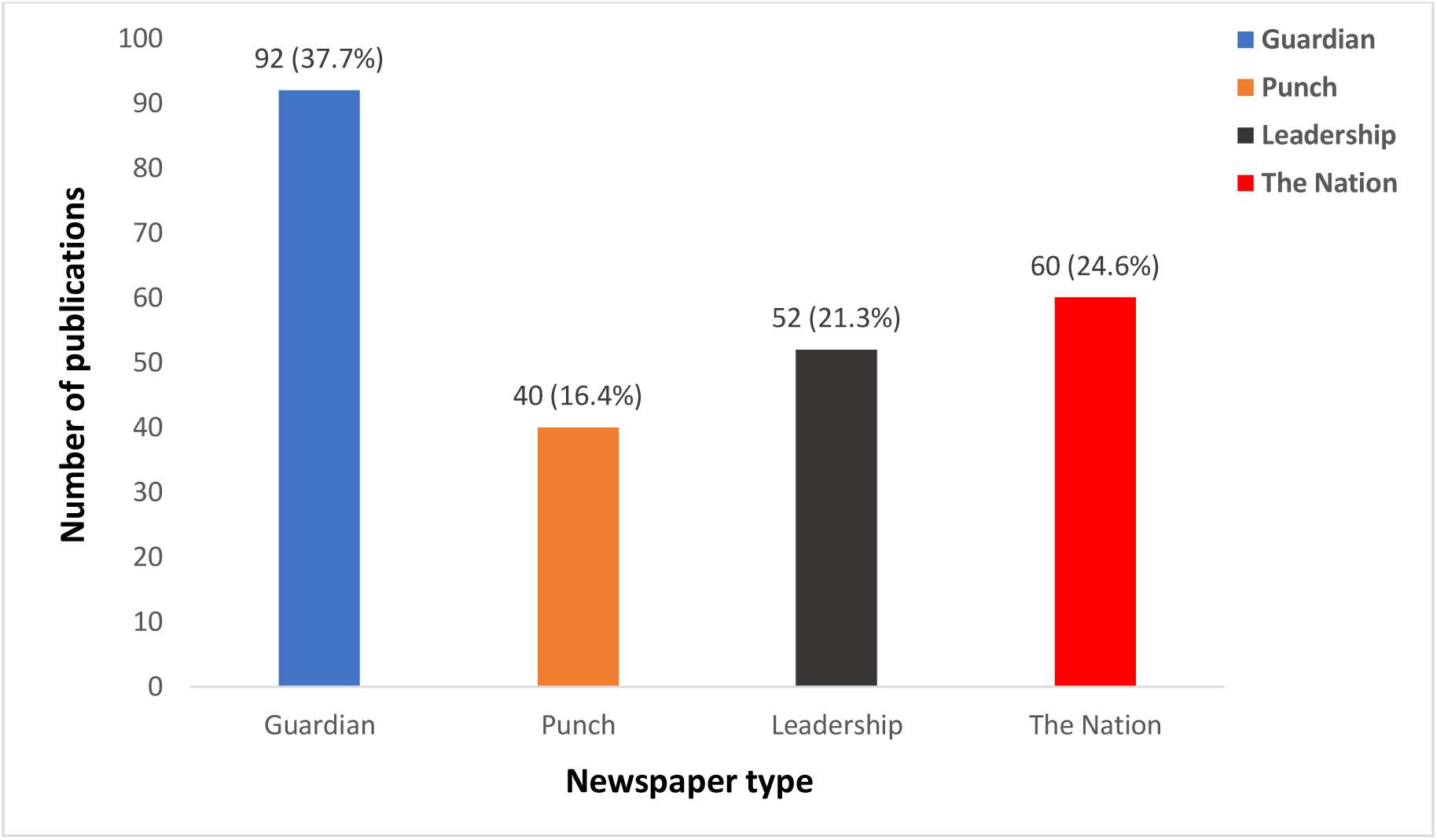
Total Publication per newspaper throughout study duration

**Table 2:**
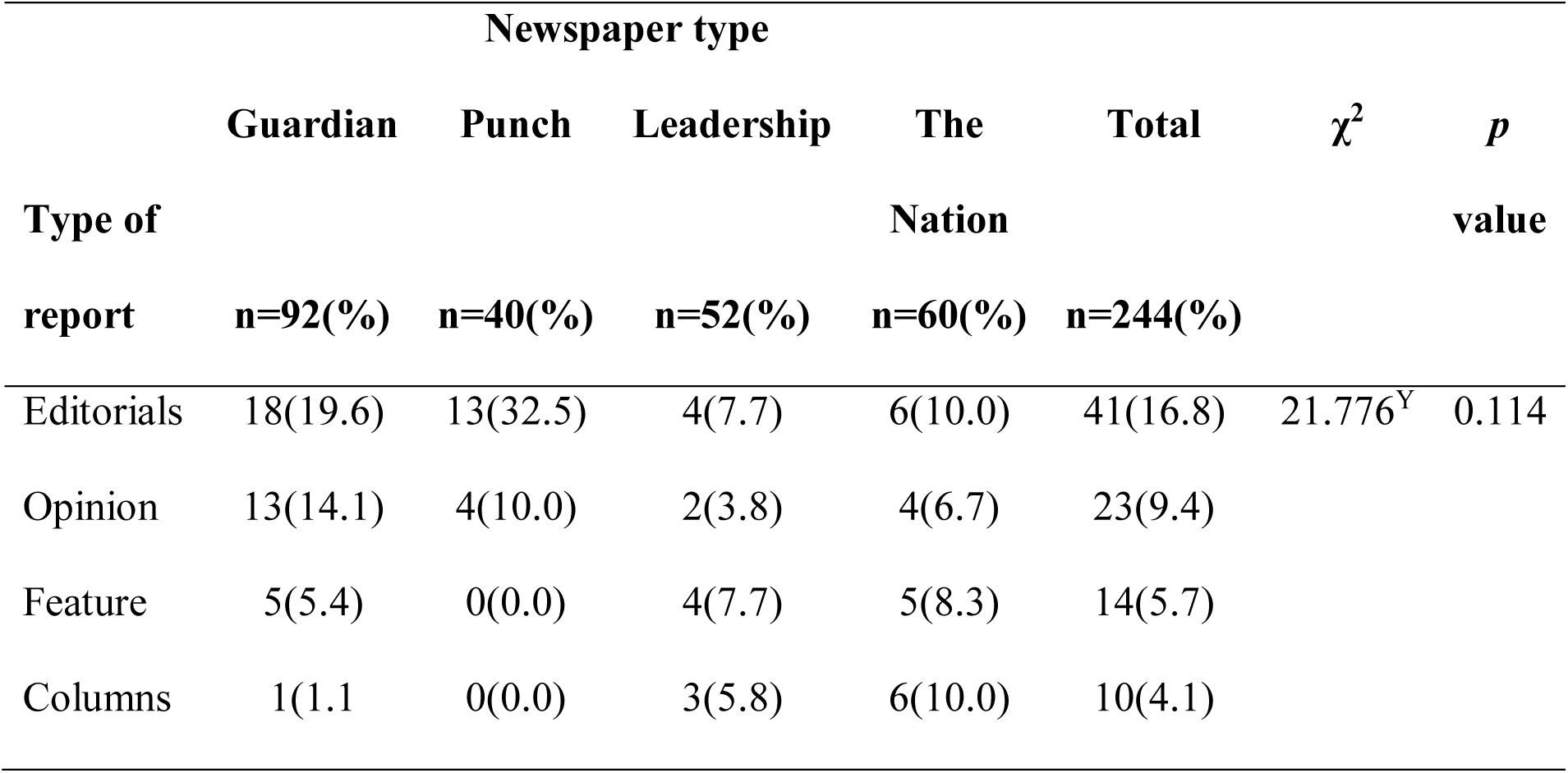
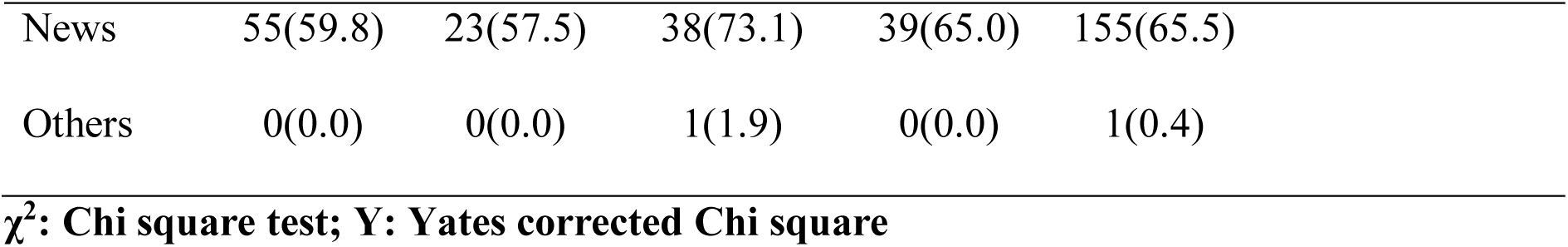
Total type of report per newspaper throughout study

| Type of report | Newspaper type | | | | | $\chi^2$ | *p* value |
| --- | --- | --- | --- | --- | --- | --- | --- |
|  | Guardian | Punch | Leadership | The Nation | Total |  |  |
|  | n=92(%) | n=40(%) | n=52(%) | n=60(%) | n=244(%) |  |  |
| Editorials | 18(19.6) | 13(32.5) | 4(7.7) | 6(10.0) | 41(16.8) | 21.776<sup>Y</sup> | 0.114 |
| Opinion | 13(14.1) | 4(10.0) | 2(3.8) | 4(6.7) | 23(9.4) |  |  |
| Feature | 5(5.4) | 0(0.0) | 4(7.7) | 5(8.3) | 14(5.7) |  |  |
| Columns | 1(1.1) | 0(0.0) | 3(5.8) | 6(10.0) | 10(4.1) |  |  |

|  |  |  |  |  |  |
| --- | --- | --- | --- | --- | --- |
| News | 55(59.8) | 23(57.5) | 38(73.1) | 39(65.0) | 155(65.5) |
| Others | 0(0.0) | 0(0.0) | 1(1.9) | 0(0.0) | 1(0.4) |

$\chi^2$ : Chi square test; Y: Yates corrected Chi square

A larger percentage of the articles appeared as news 65.5%(n=155) with 16.8%(n=41) of it also appearing as Editorials and 9.4%(n=23) appearing as opinions. 0.4%(n=1) was classified as others and this came from leadership newspaper.

A monthly breakdown of the articles was also needed to determine the frequency of reporting these stories in relation to the time difference from when the vaccine was first introduced and from the results the attention was loudest at the time of the vaccine introduction to just about a month after the introduction with the subsequent month seeing a decline as it relates to the coverage of the information.

**Table 3a:**
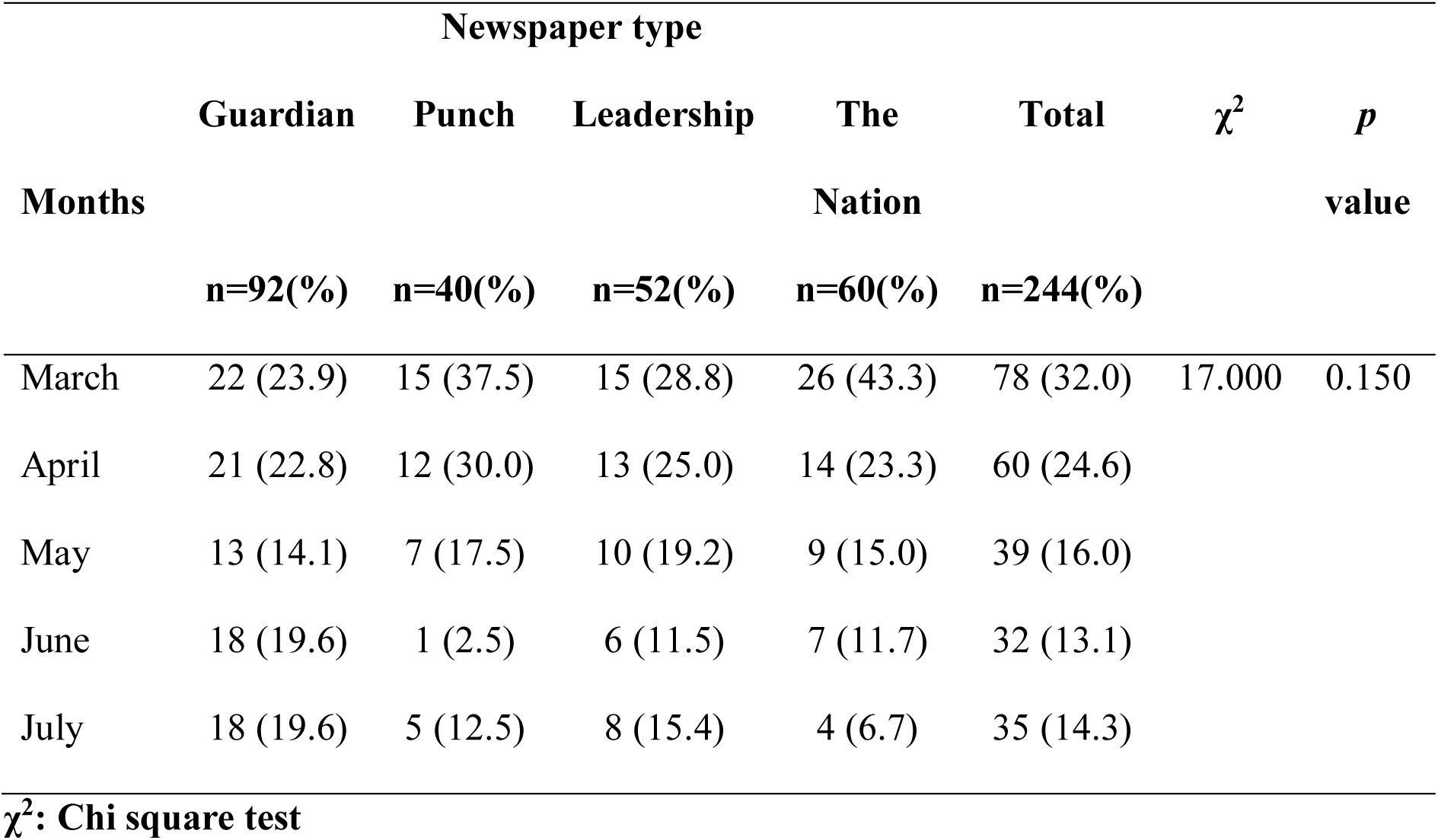
Monthly breakdown of publication

**Table 3b:** Distribution of source of information for each newspaper throughout study

| Source of information | Newspaper type | | | | | $\chi^2$ | *p* value |
| --- | --- | --- | --- | --- | --- | --- | --- |
|  | Guardian<br>n=92(%) | Punch<br>n=40(%) | Leadership<br>n=52(%) | The Nation<br>n=60(%) | Total<br>n=244(%) |  |  |
| Government Officials | 35(38.0) | 17(42.5) | 21(40.4) | 24(40.0) | 97(39.8) | 11.227<sup>Y</sup> | 0.510 |
| Technical Experts | 38(41.3) | 14(35.0) | 19(36.5) | 12(20.0) | 83(34.0) |  |  |
| Religious bodies/organizations | 3(3.3) | 4(10.0) | 4(7.7) | 9(15.0) | 20(8.2) |  |  |
| Victims/Patients | 5(5.4) | 0(0.0) | 2(3.8) | 4(6.7) | 11(4.5) |  |  |
| Others (including the newspaper) | 11(12.0) | 5(12.5) | 6(11.5) | 11(18.3) | 33(13.5) |  |  |

$\chi^2$ : Chi square test; Y: Yates corrected Chi square

Much emphasis on the information was around march accounting for 32.0% (n=78) as against what was recorded in subsequent months: April 24.6% (n=60), May 16.0% (n=39) June 13.1% (n=32) and July 14.3%(n=35). It is imperative to note that reporting in July was higher than reporting in June and this might be due to more noise around winding up of the first phase of the vaccination. The major source of information for the newspapers were Government Officials accounting for 39.8% (n=97) and Technical experts 34.0% (n=83). About 4.5% (n=11) of the information came from people who did receive the vaccines.

We also did look at the total number of articles that talked about the side effects of the COVID-19 vaccines as against those that did not mention the side effects including the various types of the side effects mentioned across the newspapers

**Table 4:**
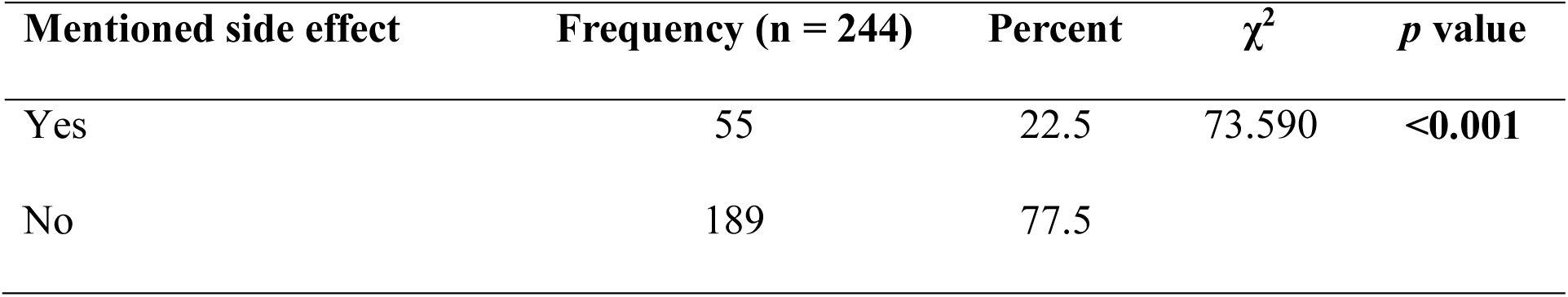
Highlights articles mentioning the vaccine side effects as against those that did not

| Mentioned side effect | Frequency (n = 244) | Percent | $\chi^2$ | *p* value |
| --- | --- | --- | --- | --- |
| Yes | 55 | 22.5 | 73.590 | <0.001 |
| No | 189 | 77.5 |  |  |

**Figure 4:**
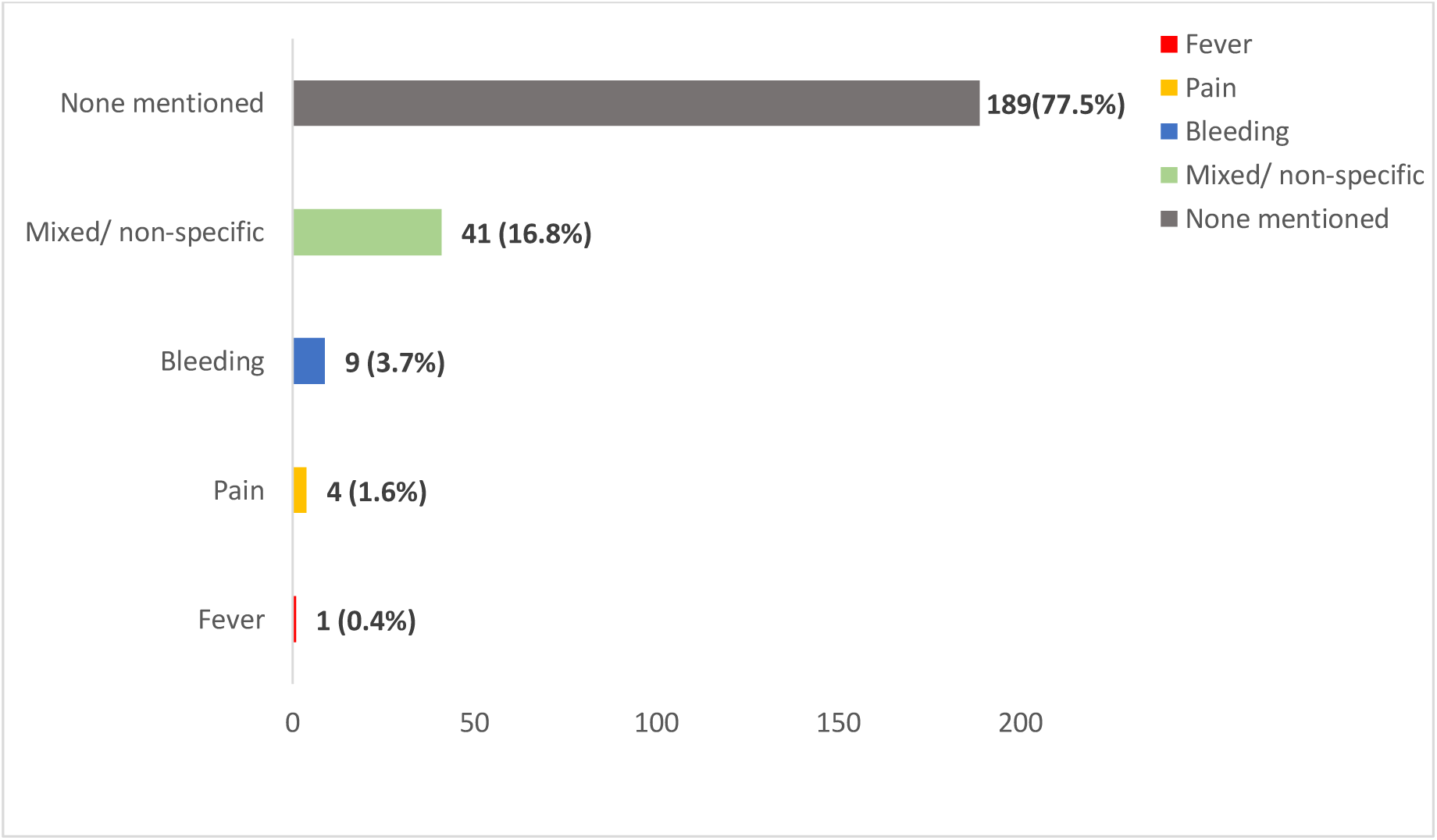
Type of side effects mentioned across the newspaper

**Table 5:** Media report on side effect of COVID-19 vaccine and where to report side effects

| Variable | Frequency (n = 244) | Percent | $\chi^2$ | *p* value |
| --- | --- | --- | --- | --- |
| **Mentioned side effect** |  |  |  |  |
| Yes | 55 | 22.5 | 73.590 | <0.001 |
| No | 189 | 77.5 |  |  |
| **Talked on where to report side effects** |  |  |  |  |
| Yes | 24 | 9.8 | 157.443 | <0.001 |
| No | 220 | 90.2 |  |  |

Majority of the newspaper 77.5% (n=189%) did not mention anything related to the side effects of the vaccine with just 22.5% (n=55). Of the number that mentioned the side effects,74.5% (n=41%) were mixed symptoms (a combination of 2 or more symptoms) while 16.4% (n=9) accounts for bleeding and 7.3%(n=4) for pain.

**Table 5 and 6** highlights those that reported the COVID-19 Vaccine side effects as well as mentioning where to report the side effects. While 22.5% (n=55) did talk about the side effects, only 9.8%(n=24) of the news article did mention where to report the side effect. Despite mentioning side effects, there was a significant difference between those that mentioned where to report the side effects as against those who mentioned side effects but not where to report the side effects with a significant level of association related to reporting the side effects.

**Table 5:** Specific side effects mentioned among those who mentioned side effects (n=55)

| Side effects mentioned | Frequency (n = 55) | Percent |
| --- | --- | --- |
| Pain | 4 | 7.3 |
| Fever | 1 | 1.8 |
| Bleeding | 9 | 16.4 |
| Mixed/ non-specific | 41 | 74.5 |

**Table 6:** Comparing newspaper that mentioned side effect of covid-19 vaccine and those who talked on where to report the side effects

| Talked on where to<br>report side effects | Mentioned side effect | | Total<br>N | $\chi^2$ | *p* value |
| --- | --- | --- | --- | --- | --- |
|  | Yes<br>n (%) | No<br>n (%) |  |  |  |
| Yes | 24 (100.0) | 0 (0.0) | 24 | 91.470 | <0.001 |
| No | 31 (14.1) | 189 (85.9) | 220 |  |  |
| Total | 55 (22.5) | 189 (77.5) | 244 |  |  |

$\chi^2$ : Chi square test

## Discussion

Our study looked at the media coverage of COVID-19 vaccine in Nigeria newspaper starting from when the vaccine was first introduced into the country (March 20210 to end of first round of the vaccination completion (July 2021) with a concentrated focus on what the source of information for the newspaper articles as well as how they talked about the side effects of the vaccine and where to report any side effect especially for those that have taken the vaccine. With a total of 244 articles reviewed, a higher frequency of coverage was seen in March 2021 (32.0%) which was the month when the vaccine was first introduced in the country with a subsequent decline in media coverage across the month, this is not unusual as Falagas et.al^20^ in their findings in evaluating the role of media during the Influenza pandemic in Greece had described 2 states; A state of alarm in October 2005 when there was extensive media coverage of the disease prompting increase vaccine uptake and a state of Silence in November /December 2005 leading to poor uptake of the vaccine. It is also important to point at Zhou et. al^21^ findings in the role the media can play in curtailing the disease spread in relation to setting an agenda and the frame to which they report the disease outbreak

During a disease outbreak, the source of information is of high importance and the information must not only be factual, reliable but also accurate with no elements of deceit. Most times, the journalist approach the technical experts to hear their views in other for them to come up with a fact -backed up story.^22^ In our study, Political office holders were the predominant major source of information accounting for 39.8% (n=970 followed by Technical Experts accounting for 34.0%(n=83). This could be as a result of the government talking more to the media as a way of trying to instil confidence in the people to take the vaccine as well as trying to be seen working hard in curtailing the spread of the disease in the country. Few information did come from those that had received the vaccines 4.5% (n=11) and we strongly feel this could have been looked more into as their first-hand experience could help sway the increase in uptake of the vaccine^23^.

While 22.5%(n=55) of the total story did talk about the side effects of the vaccines, the reported the side effects in a mixed way and not as individual entities (More than 1 side effects accounts for 74.5% of the total n=55) while bleeding was reported most of the single entity accounting for 16.4% of the side effects mentioned. This is not so as the very common side effects listed in the prescribing information leaflet of AstraZeneca where injection site pain, fever, headache and myalgia were listed as the commonest side effects^24^ One reason that could explain why lots of the emphasis on the bleeding tendency could be on the attention that this side effect of the vaccine got on a global scale necessitating some countries withholding the administration of the vaccine as well as some countries switching to other type of the vaccines ^25^

Talking about where to report the side effects of the vaccines to people who have had the vaccine administered would not only help capture the data on the side effects but would also serve as a booster for them showing some level of care. Our study showed a statistical significance between the articles that mentioned the side effects and where the people are to report the side effects (p-value <0.001).

This study to the best of our knowledge and search is the only one which has focused entirely on this topic in the country and it further buttresses the need for continuous campaign about the role of the media in helping to promote the benefits as well as the risk of the COVID-19 vaccine in the country including pointing he citizenry in the right direction if they need to report any side effects following administration of the vaccine.

## Conclusions

Our conclusion is that coverage of the COVID-19 vaccine administration including the side effects in the country by the print media is still inadequate considering the hesitancy and the palpable fears among people about the vaccine. The source of the information used by the newspaper are good however they can also focus more on the perspective of recipients of the vaccines and the various outcomes they experienced after taking the vaccine. While side effects are not reported in the newspaper and the regulatory bodies had provided information on where to report the side effects for anyone who took the vaccines, the print media did not provide much information on where to report the side effects and how to report it.

We strongly believe that as part of the strategies to help increase the uptake of the vaccines across the country, the media has a significant role in preaching the right message to the wider community. Future research might want to expand the scope of this research as well as expand the timeline to see if the situation has changed.

## Limitations

Only 4 national newspapers were sampled out of the over 30 national newspapers in the country and we might have inadvertently excluded some with wider coverage of the topic. We also recognised that some other form of media such as television, radio and billboards might cover this topic in a more extensive way than the print media.

## Data Availability

Availability of Data and Materials
The data used and or analysed in this study is available on request from the corresponding author on reasonable request.

## Availability of Data and Materials

The data used and or analysed in this study is available on request from the corresponding author on reasonable request.

